# The Impact of Social Determinants of Health and Lifestyle on the Association of Lipoprotein(a) with Myocardial Infarction and Stroke

**DOI:** 10.1101/2023.09.01.23294968

**Authors:** Eric J. Brandt, Matthias Kirch, Nimai Patel, Chaitanya Chennareddy, Venkatesh L. Murthy, Sascha N. Goonewardena

## Abstract

**Background:** In European cohorts, a higher Mediterranean diet or Life’s Simple 7 (LS7) score abolished or attenuated the risk associated with increasing Lipoprotein(a) [Lp(a)] on cardiovascular outcomes. This is unstudied in US cohorts. The impact of social determinants of health (SDOH) on the association of Lp(a) with cardiovascular outcomes remains unstudied. We sought to test if a SDOH score and LS7 score impacts the association of Lp(a) with myocardial infarction (MI) or stroke.

**Methods:** Observational Cohort of US Adults from the Atherosclerosis Risk in Communities (ARIC) and Multi-Ethnic Study of Atherosclerosis (MESA) cohorts. We performed sequential multivariable Cox proportional hazard analysis, first adjusting for age, gender, non-HDL-C, race and ethnicity, then added SDOH and LS7 scores sequentially. The primary outcomes were time until first fatal or nonfatal MI or stroke.

**Results:** ARIC (n=15,072; median Lp(a)=17.3 mg/dL) had 16.2 years average follow up. MESA (n=6,822; median Lp(a)=18.3 mg/dL had 12.3 years average follow-up. In multivariable analyses adjusted for age, gender, race and ethnicity, and non-HDL-C, Lp(a) was associated (HR, p-value) with MI in ARIC (1.10, <0.001) and MESA (1.09, <0.001), and stroke in ARIC (1.08, <0.001) but not MESA (0.97, 0.50). With SDOH and LS7 added to the model associations remained similar (association of Lp(a) with MI in ARIC 1.09, <0.001 and in MESA 1.10, 0.001, with stroke in ARIC 1.06, <0.003 and in MESA 0.96, 0.39). In models with all covariates, each additional SDOH correlated positively with MI (ARIC 1.13, <0.001; MESA 1.11, <0.001) and stroke (ARIC 1.17, <0.001; HR 1.07, p=0.11) and each additional LS7 score point correlated negatively with MI (ARIC 0.81, <0.001; MESA 0.84, <0.001) and stroke (ARIC 0.82, <0.001; MESA 0.84, <0.001).

**Conclusions and Relevance:** SDOH and lifestyle factors were predictors for MI and stroke that did not impact the association between Lp(a) and cardiovascular events. Our findings support that Lp(a) is an independent risk factor for MI and possibly stroke.

## Introduction

Lipoprotein(a) (Lp(a)) is an independent risk factor for the development of atherosclerotic cardiovascular diseases (ASCVD).^1^ Aside from apheresis, there are no clinically available therapies that specifically target lowering of Lp(a), although agents are being developed (e.g., pelacarsen, olpasiran, etc.) and other FDA-approved therapies (e.g., PCSK9 inhibitors and Niacin) have small impacts on Lp(a) levels. While awaiting therapeutic agents, patients with elevated Lp(a) can lower their risk for ASCVD with statins and other lipids lowering therapies.^1^ Additionally, lifestyle and social determinants of health (SDOH) have large impacts on cardiovascular outcomes and can be targets for risk modification.^2,3^

Findings from two European cohorts suggest that a healthy lifestyle can decrease the risk for ASCVD among those with elevated Lp(a). In the EPIC-Norfolk cohort (n=14,051 men and women from Norfolk, UK), those in the top versus the bottom tertile of Life’s Simple 7 (LS7) score and Lp(a) ≥50 mg/dL had a hazard ratio (HR) of 0.33 (95% CI, 0.17-0.63) for death from coronary heart disease or stroke.^4^ The risk of elevated Lp(a) was not completely abolished since those with Lp(a) <50 mg/dL in the top tertile of LS7 score had a HR of 0.17 (95% CI 0.12-0.31). Second, in the ATTICA study (n=3,042 Greek men and women) without ASCVD, a higher Mediterranean diet score did not influence Lp(a) levels, but did abolish (HR 1.00, 95%CI 0.98-1.01) the risk associated with Lp(a) for having an ASCVD event.^5^

SDOH are the conditions in which people are born, live, and work.^6^ SDOH are associated with ASCVD events and may help to explain the differences in cardiovascular risk between different populations.^7–9^ SDOH can impact risk for ASCVD via psychological, behavioral, and biologic mechanisms, which include chronic stress responses and systemic inflammation.^10^ Given that Lp(a) is an acute phase reactant whose level is mediated by periods of inflammation, it is plausible that SDOH could impact the association of Lp(a) on risk for CVD events.^11^

The impact of lifestyle factors on the association of Lp(a) with cardiovascular events has not been tested in US cohorts. Further, the impact of SDOH on the association of Lp(a) with cardiovascular outcomes is untested in any cohorts. We hypothesized that accounting for SDOH and lifestyle factors would greatly mitigate the risk associated with Lp(a) for myocardial infarction (MI) or stroke in US cohorts.

## Methods

### Data

We performed retrospective analysis of prospectively data from the Atherosclerosis Risk in Communities (ARIC) and the Multi-Ethnic Study of Atherosclerosis (MESA) cohorts. ARIC and MESA are prospective, longitudinal investigations into the cardiac risk factors, health outcomes, and demographic patterning of atherosclerosis. ARIC enrolled men and women aged 35-84 years, beginning in 1987 from four US communities. MESA enrolled men and women aged 45-64 years, beginning in 2000 from six US communities. Data were obtained via requests to the National Institute of Health’s Biologic Specimen and Data Repository Information Coordinating Center. We used data from the first examination cycle and cohort surveillance data for cardiovascular events. The study was deemed exempt from review by the University of Michigan institutional review board.

### Outcomes

The primary outcomes were time until first fatal or nonfatal MI or stroke. In ARIC, outcomes were tracked semi-annually through 2006. In MESA, outcomes were tracked annually through 2015.

### Lipoprotein(a) assays

In ARIC, Lp(a) was measured as protein mass using a double-antibody enzyme-linked immunosorbent assay technique.^12,13^ The protein-mass represents about one-third of the total molecule mass,^14^ thus the Lp(a)-mass measured in ARIC Visit 1 was tripled to be similar to the Lp(a) total-mass measurement in MESA. This technique, although isoform sensitive, had excellent correlation (r=0.88) with samples performed at Visit 4 using an isoform insensitive turbidimetric immunoassay (Denka Seiken Co. Ltd., Tokyo, Japan).^15^ In MESA, Lp(a) was measured from cold storage 10-11 years after sample collection as mass content using an isoform insensitive latex-enhanced turbidimetric immunoassay (Denka Seiken, Tokyo, Japan) by Health Diagnostics Laboratory (Richmond, Virginia).^16,17^

### Statistical Design and Analysis

We performed sequential multivariable cox proportional hazard analysis. In Model 1, covariates were age, gender, race and ethnicity, non-HDL-C (per 25 mg/dL increase), and Lp(a) (per 25 mg/dL increase). Model 2 included a SDOH score: an integer score ranging from zero to five in ARIC and zero to 11 in MESA. SDOH in ARIC included being unemployed, income <300% of the federal poverty level, <high school education, no regular site for healthcare access, and government or no health insurance. SDOH in MESA included these determinants and not being married or living with a partner, not owning a home, reports of loneliness/lack of social support, unsafe neighborhood residence, experience of discrimination in the last year, and somewhat or very serious food access problems. Model 3 included the LS7 score (smoking status, body mass index, physical activity, dietary score, total cholesterol, blood pressure, and fasting plasma glucose), which has been previously defined.^3^ Each category received zero to two points (range 0-14). Non-HDL-C was calculated by subtracting HDL-C from Total Cholesterol and corrected for Lp(a) mass (non-HDL-C=Total Cholesterol–HDL-C–(Lp(a)x0.3)).^18^ We used interaction terms to test whether Lp(a) is moderated by SDOH or LS7 scores.

To test the proportional hazards assumption we examined log-log plots of survival for parallel curves in all fully constructed models, which were acceptable for all outcomes. All p-values were two-sided. Statistical significance was set at p<0.05. Data were analyzed using Stata software, version 16 (StataCorp, LLC).

### Sensitivity analyses

To understand if correlations between SDOH and LS7 scores could impact outcomes, we tested whether SDOH or LS7 scores predicted Lp(a) level in age, gender, race and ethnicity, and non-HDL-C adjusted models.

To understand the association between categorical Lp(a) level and cardiovascular outcomes we use coarsened exact matching to match cases (Lp(a) >50 mg/dL) to controls (Lp(a) <50 mg/dL). In ARIC, matching covariates were coarsened to age (44-55, or 56-66), non-HDL-C (0-120.0, 120.1-160.0, 160.1-200.0, 200.1-240.0, and ≥240.1 mg/dL), SDOH score (0, 1-2, 3-5), and LS7 score (0-4, 5-9, 10-14). Non-coarsened variables included gender and race and ethnicity. In MESA matching covariates were similar except age (44-57, 57-70, and 70-84) and SDOH score (0-1, 1-3, 3-5, >5). We repeated cox proportional hazards regression in the same three models as described above except that we consider Lp(a) as a three-level outcome (<50 mg/dL, ≥50 to <100 mg/dL, and ≥100 mg/dL).

## Results

### Population Characteristics

In ARIC the mean age was 54 years and in MESA 62 years. Most were female (54.5% in ARIC and 52.9% in MESA, **Table 1**). The largest race and ethnicity represented was non-Hispanic White (73.9% in ARIC, 38.5% in MESA), followed by non-Hispanic Black (26.1% in ARIC, 27.8% in MESA), and only MESA included Chinese (11.8%) and Hispanic (22.0%) individuals. Mean Lp(a) was similar in both cohorts (30.3 mg/dL in ARIC and 30.1 mg/dL in MESA), whereas non-HDL-C was higher in ARIC (155 mg/dL) than MESA (134 mg/dL). SDOH scores were lower in ARIC (mean(SD)=1.0(1.1)) than MESA (mean=2.3(1.7)). Mean(SD) LS7 score was similar in ARIC (7.8(2.3)) and MESA (8.3(2.1)).

**Table 1:** Population characteristics (n=15,027 for ARIC, n=6,822 for MESA)

|  | ARIC<br>n, % or mean (SD) | MESA<br>n, % or mean (SD) |
| --- | --- | --- |
| Age (years) | 54 (6) | 62 (10) |
| Female | 8196, 54.5% | 3601, 52.9% |
| Race and Ethnicity | - | - |
| Non-Hispanic White | 11,110, 73.9% | 2,622, 38.5% |
| Non-Hispanic Black | 3917, 26.1% | 1892, 27.8% |
| Non-Hispanic Chinese | n/a | 804, 11.8% |
| Hispanic | n/a | 1496, 22.0% |
| Lipoprotein(a) (mg/dL) | 30.3 (32.1) | 30.1 (34.0) |
| Non-HDL-C (mg/dL) | 155 (43.9) | 134 (36.1) |
| SDOH Score | 1.0 (1.1) | 2.3 (1.7) |
| Life's Simple 7 Score | 7.8 (2.3) | 8.3 (2.1) |

Abbreviations: ARIC = Atherosclerosis Risk in Communities; MESA=Multi-Ethnic Study of Atherosclerosis; Non- HDL-C = Non-High Density Lipoprotein-Cholesterol; SDOH = Social Determinants of Health

### Lp(a) association with SDOH and LS7 scores

In age, gender, and race and ethnicity adjusted models, SDOH score did not predict Lp(a) level (increase in mg/dL Lp(a) per SDOH), 95% CI) in either ARIC (0.50 (-0.003, 1.00)) or MESA (0.24 (-0.27, 0.74)). However, LS7 score did predict lower Lp(a) level in both ARIC (-0.68 (-0.90, -0.45) and MESA (-0.46, (-0.85, -0.07)).

### Myocardial Infarction

In all models, Lp(a) (per 25 mg/dL increase) was associated with MI (**Table 2**). The HR (95% CI) only slightly attenuated after adding SDOH and LS7 into the models (in ARIC from 1.11 (1.08, 1.14) to 1.09 (1.06, 11) and in MESA from 1.11 (1.05, 1.17) to 1.10 (1.04, 1.16)). The association between non-HDL-C and MI in ARIC (1.16 (1.14, 1.18)) and MESA (1.09 (1.03, 1.05)) was similar when adding SDOH (1.15 (1.13, 1.17) in ARIC and 1.09 (1.03, 1.15) in MESA), then attenuated substantially once LS7 score was added in ARIC (1.04 (1.02, 1.06) and nonsignificant in MESA (1.02 (0.96, 1.08)).

**Table 2:** Sequential Multivariable Cox Proportional Hazards Regression for Myocardial Infarction in ARIC and MESA.

A: ARIC (n=14,306)
|  | Model 1 |  |  | Model 2 |  |  | Model 3 |  |  |
| --- | --- | --- | --- | --- | --- | --- | --- | --- | --- |
|  | HR | 95% CI | p-value | HR | 95% CI | p-value | HR | 95% CI | p-value |
| Age (per year) | 1.09 | 1.09, 1.10 | <0.001 | 1.09 | 1.08, 1.10 | <0.001 | 1.09 | 1.08, 1.10 | <0.001 |
| Male | 1.62 | 1.52, 1.74 | <0.001 | 1.62 | 1.52, 1.73 | <0.001 | 1.60 | 1.50, 1.71 | <0.001 |
| Race and Ethnicity | - | - | - | - | - | - | - | - | - |
| Non-Hispanic White | Ref | Ref | Ref | Ref | Ref | Ref | Ref | Ref | Ref |
| Non-Hispanic Black | 1.34 | 1.24, 1.45 | <0.001 | 1.13 | 1.04, 1.23 | 0.006 | 0.88 | 0.80, 0.96 | 0.003 |
| Non-HDL-C (per 25 mg/dL increase) | 1.16 | 1.14, 1.18 | <0.001 | 1.15 | 1.13, 1.17 | <0.001 | 1.04 | 1.02, 1.06 | <0.001 |
| Lipoprotein(a) (per 25 mg/dL increase) | 1.11 | 1.08, 1.14 | <0.001 | 1.11 | 1.08, 1.14 | <0.001 | 1.09 | 1.06, 1.12 | <0.001 |
| SDOH Score (per 1 point increase) | - | - | - | 1.22 | 1.18, 1.26 | <0.001 | 1.13 | 1.09, 1.17 | <0.001 |
| Life's Simple 7 Score (per 1 point increase) | - | - | - | - | - | - | 0.81 | 0.80, 0.83 | <0.001 |

B: MESA (n=6,668)
|  | Model 1 |  |  | Model 2 |  |  | Model 3 |  |  |
| --- | --- | --- | --- | --- | --- | --- | --- | --- | --- |
|  | HR | 95% CI | p-value | HR | 95% CI | p-value | HR | 95% CI | p-value |
| Age (per year) | 1.06 | 1.05, 1.07 | <0.001 | 1.05 | 1.04, 1.06 | <0.001 | 1.05 | 1.04, 1.06 | <0.001 |
| Male | 2.19 | 1.85, 2.58 | <0.001 | 2.36 | 2.00, 2.79 | <0.001 | 2.37 | 2.00, 2.80 | <0.001 |
| Race and Ethnicity | - | - | - | - | - | - | - | - | - |
| Non-Hispanic White | Ref | Ref | Ref | Ref | Ref | Ref | Ref | Ref | Ref |
| Non-Hispanic Black | 0.95 | 0.78, 1.17 | 0.65 | 0.86 | 0.70, 1.06 | 0.16 | 0.72 | 0.58, 0.89 | 0.002 |
| Non-Hispanic Chinese | 0.78 | 0.59, 1.03 | 0.08 | 0.69 | 0.52, 0.92 | 0.01 | 0.78 | 0.58, 1.03 | 0.08 |
| Hispanic | 1.01 | 0.82, 1.24 | 0.92 | 0.81 | 0.65, 1.02 | 0.07 | 0.75 | 0.59, 0.94 | 0.012 |
| Non-HDL-C (per 25 mg/dL increase) | 1.09 | 1.03, 1.15 | 0.002 | 1.09 | 1.03, 1.15 | 0.003 | 1.02 | 0.96, 1.08 | 0.53 |
| Lipoprotein(a) (per 25 mg/dL increase) | 1.11 | 1.05, 1.17 | <0.001 | 1.11 | 1.05, 1.17 | <0.001 | 1.10 | 1.04, 1.16 | 0.001 |
| SDOH Score (per 1 point increase) | - | - | - | 1.14 | 1.08, 1.20 | <0.001 | 1.11 | 1.05, 1.17 | <0.001 |
| Life's Simple 7 Score (per 1 point increase) | - | - | - | - | - | - | 0.84 | 0.81, 0.88 | <0.001 |

Among demographic characteristics, there was no change in the association between age and MI and only a small change in association between gender and MI when adding SDOH and LS7 score (**Table 2**). There were changes in the association between race and ethnicity and MI when adding SDOH and LS7 into the models; the association of identifying as non-Hispanic Black compared to non-Hispanic White changed in ARIC from 1.34 (1.24, 1.45) to 0.88 (0.80, 0.96) and in MESA from 0.95 (0.78, 1.17) to 0.72 (0.58, 0.89). For those in MESA identifying as Hispanic, the association shifted from 1.01 (0.82, 1.24) to 0.75 (0.59, 0.94). There was no significant shift for those identifying as non-Hispanic Chinese. Given the large changes in association we also tested for interactions between race and ethnicity with SDOH and LS7 scores, all of which were nonsignificant (p>0.05).

There were no significant interactions between SDOH and Lp(a) on MI in ARIC (HR 1.01, p=0.46) or MESA (1.01, p=0.68). There was an interaction with a small effect size between LS7 and Lp(a) on MI in MESA (0.97, p=0.03), but not ARIC (1.00, p=0.58).

### Stroke

Lp(a) associated with stroke in ARIC, but not in MESA (**Table 3**). In ARIC, the association with stroke (HR (95% CI)) only slightly declined from 1.08 (1.05, 1.12) to 1.06 (1.02, 1.09) when including SDOH and LS7 scores. The association between non-HDL-C and stroke in ARIC (1.09 (1.06, 1.11)) and MESA (1.10 (1.01, 1.19) decreased slightly when adding SDOH (1.08 (1.05, 1.11) in ARIC and 1.10 (1.02, 1.19) in MESA)), then became non-significant once LS7 score was added (0.98 (0.95, 1.01) in ARIC and 1.03 (0.94, 1.12) in MESA).

**Table 3:** Sequential Multivariable Cox Proportional Hazards Regression for Stroke in ARIC and MESA A: ARIC (n=14,286)

|  | Model 1 |  |  | Model 2 |  |  | Model 3 |  |  |
| --- | --- | --- | --- | --- | --- | --- | --- | --- | --- |
|  | HR | 95% CI | p-value | HR | 95% CI | p-value | HR | 95% CI | p-value |
| Age (per year) | 1.11 | 1.10, 1.12 | <0.001 | 1.11 | 1.10, 1.12 | <0.001 | 1.11 | 1.10, 1.12 | <0.001 |
| Male | 1.28 | 1.16, 1.40 | <0.001 | 1.27 | 1.15, 1.39 | <0.001 | 1.25 | 1.14, 1.38 | <0.001 |
| Race and Ethnicity | - | - | - | - | - | - | - | - | - |
| Non-Hispanic White | Ref | Ref | Ref | Ref | Ref | Ref | Ref | Ref | Ref |
| Non-Hispanic Black | 1.96 | 1.77, 2.18 | <0.001 | 1.60 | 1.43, 1.80 | <0.001 | 1.24 | 1.10, 1.40 | <0.001 |
| Non-HDL-C (per 25 mg/dL increase) | 1.09 | 1.06, 1.11 | <0.001 | 1.08 | 1.05, 1.11 | <0.001 | 0.98 | 0.95, 1.01 | 0.13 |
| Lipoprotein(a) (per 25 mg/dL increase) | 1.08 | 1.05, 1.12 | <0.001 | 1.08 | 1.04, 1.12 | <0.001 | 1.06 | 1.02, 1.09 | 0.003 |
| SDOH Score (per 1 point increase) | - | - | - | 1.26 | 1.20, 1.31 | <0.001 | 1.17 | 1.12, 1.22 | <0.001 |
| Life's Simple 7 Score (per 1 point increase) | - | - | - | - | - | - | 0.82 | 0.80, 0.84 | <0.001 |

B: MESA (n=6,666)
|  | Model 1 |  |  | Model 2 |  |  | Model 3 |  |  |
| --- | --- | --- | --- | --- | --- | --- | --- | --- | --- |
|  | HR | 95% CI | p-value | HR | 95% CI | p-value | HR | 95% CI | p-value |
| Age (per year) | 1.07 | 1.06, 1.09 | <0.001 | 1.07 | 1.06, 1.08 | <0.001 | 1.07 | 1.06, 1.08 | <0.001 |
| Male | 1.26 | 1.00, 1.59 | 0.05 | 1.34 | 1.05, 1.70 | 0.02 | 1.34 | 1.05, 1.70 | 0.02 |
| Race and Ethnicity | - | - | - | - | - | - | - | - | - |
| Non-Hispanic White | Ref | Ref | Ref | Ref | Ref | Ref | Ref | Ref | Ref |
| Non-Hispanic Black | 1.43 | 1.06, 1.95 | 0.02 | 1.34 | 0.99, 1.83 | 0.06 | 1.12 | 0.82, 1.54 | 0.47 |
| Non-Hispanic Chinese | 0.64 | 0.40, 1.04 | 0.07 | 0.59 | 0.36, 0.96 | 0.03 | 0.66 | 0.41, 1.08 | 0.10 |
| Hispanic | 1.53 | 1.14, 2.06 | 0.005 | 1.31 | 0.95, 1.82 | 0.10 | 1.20 | 0.87, 1.66 | 0.27 |
| Non-HDL-C (per 25 mg/dL increase) | 1.10 | 1.01, 1.19 | 0.02 | 1.10 | 1.01, 1.19 | 0.03 | 1.03 | 0.94, 1.12 | 0.54 |
| Lipoprotein(a) (per 25 mg/dL increase) | 0.97 | 0.89, 1.07 | 0.58 | 0.97 | 0.88, 1.07 | 0.56 | 0.96 | 0.87, 1.05 | 0.39 |
| SDOH Score (per 1 point increase) | - | - | - | 1.10 | 1.02, 1.19 | 0.02 | 1.07 | 0.99, 1.15 | 0.11 |
| Life's Simple 7 Score (per 1 point increase) | - | - | - | - | - | - | 0.84 | 0.79, 0.89 | <0.001 |

Among demographic characteristics, there was no change in association between age and stroke and only a slight shift between gender and stroke when adding SDOH and LS7 score to the models (**Table 3**). Again there were shifts in associations for race and ethnicity and stroke; identifying as non-Hispanic Black compared to non-Hispanic White changed when adding SDOH and LS7 scores to the models in ARIC from HR 1.96 (1.77, 2.18) to 1.24 (1.10, 1.40) and in MESA from 1.43 (1.06, 1.95) to 1.12 (0.82, 1.54). For those in MESA identifying as Hispanic, the association also shifted from 1.53 (1.14, 2.06) to 1.20 (0.87, 1.66). There was no significant shift for those identifying as non-Hispanic Chinese. Given the large changes in association we also tested for interactions between race and ethnicity with SDOH and LS7 scores, all of which were nonsignificant (p>0.05).

There were no significant interactions between SDOH and Lp(a) on stroke in ARIC (HR 1.01, p=0.38) or MESA (1.01, 0.67) nor between LS7 and Lp(a) on stroke in ARIC (0.99, 0.40) or MESA (0.98, 0.36).

### Coarsened Exact Matching

In ARIC, there were 214 (1.9% of those with Lp(a) <50 mg/dL) individuals unmatched with Lp(a) <50 mg/dL and 535 (14.8%) with Lp(a) ≥50 mg/dL. In MESA, there were 1004 (18.7%) individuals unmatched with Lp(a) <50 mg/dL and 146 (10.0%) with Lp(a) >50 mg/dL. In ARIC, weight cases were similar to weighted controls in all factors except for Lp(a) (81.3 mg/dL vs 19.4 mg/dL, p<0.001) (**Table 4**). In MESA, cases were similar to controls except for non-HDL-C (122.7 mg/dL vs 125.8 mg/dL, p=0.005) and Lp(a) (86.0 mg/dL vs 18.8 mg/dL, p<0.001).

**Table 4:** Population Characteristics Cases with Lipoprotein(a) ≥50 mg/dL and Controls with Lipoprotein(a) <50 mg/dL after Coarsened Exact Matching Weights Applied.

A: ARIC
|  | Controls<br>n, % or mean (SD) | Cases<br>n, % or mean (SD) | p-value |
| --- | --- | --- | --- |
| Age (years) | 54.3 (5.8) | 54.3 (5.8) | 0.66 |
| Female | 6826, 60.9% | 1872, 60.9% | 1.00 |
| Race and Ethnicity | - | - | - |
| Non-Hispanic White | 6224, 44.5% | 1366, 44.5% | 1.00 |
| Non-Hispanic Black | 4981, 55.6% | 1366, 55.6% | 1.00 |
| Lipoprotein(a) (mg/dL) | 19.4 (13.9) | 81.3 (30.1) | $< 0.001$ |
| Non-HDL-C (mg/dL) | 150 (45.0) | 149 (44.4) | 0.73 |
| SDOH Score | 1.2 (1.2) | 1.2 (1.1) | 0.80 |
| Life's Simple 7 Score | 7.5 (2.3) | 7.4 (2.3) | 0.07 |

B: MESA
|  | Controls<br>n, % or mean (SD) | Cases<br>n, % or mean (SD) | p-value |
| --- | --- | --- | --- |
| Age (years) | 62.3 (9.9) | 62.3 (10.0) | 0.83 |
| Female | 2554, 5.87% | 773, 58.7% | 1.00 |
| Race and Ethnicity | - | - | - |
| Non-Hispanic White | 1364, 31.4% | 413, 31.4% | 1.00 |
| Non-Hispanic Black | 2051, 47.2% | 621, 47.2% | 1.00 |
| Non-Hispanic Chinese | 248, 5.7% | 75, 5.7% | 1.00 |
| Hispanic | 683, 15.7% | 207, 15.7% | 1.00 |
| Lipoprotein(a) (mg/dL) | 18.8 (13.3) | 86.0 (34.6) | $< 0.001$ |
| Non-HDL-C (mg/dL) | 125.8 (33.3) | 122.7 (35.3) | 0.005 |
| SDOH Score | 2.3 (1.7) | 2.3 (1.7) | 0.78 |
| Life's Simple 7 Score | 8.2 (2.0) | 8.1 (2.0) | 0.38 |

In fully adjusted models that included SDOH score and LS7 score, Lp(a) ≥50 mg/dL to <100 mg/dL was associated with MI in ARIC (HR 1.13,p <0.001) and MESA (1.12, 0.12) (**Figure 1**). Lp(a) ≥100 mg/dL was also associated with MI in ARIC (1.44, p<0.001) and MESA (1.57, 0.007). Associations between other covariates and MI were similar, including similar shifts in association between race and ethnicity with MI after adding SDOH and LS7 scores to the model (**Table 5**).

**Figure 1:**
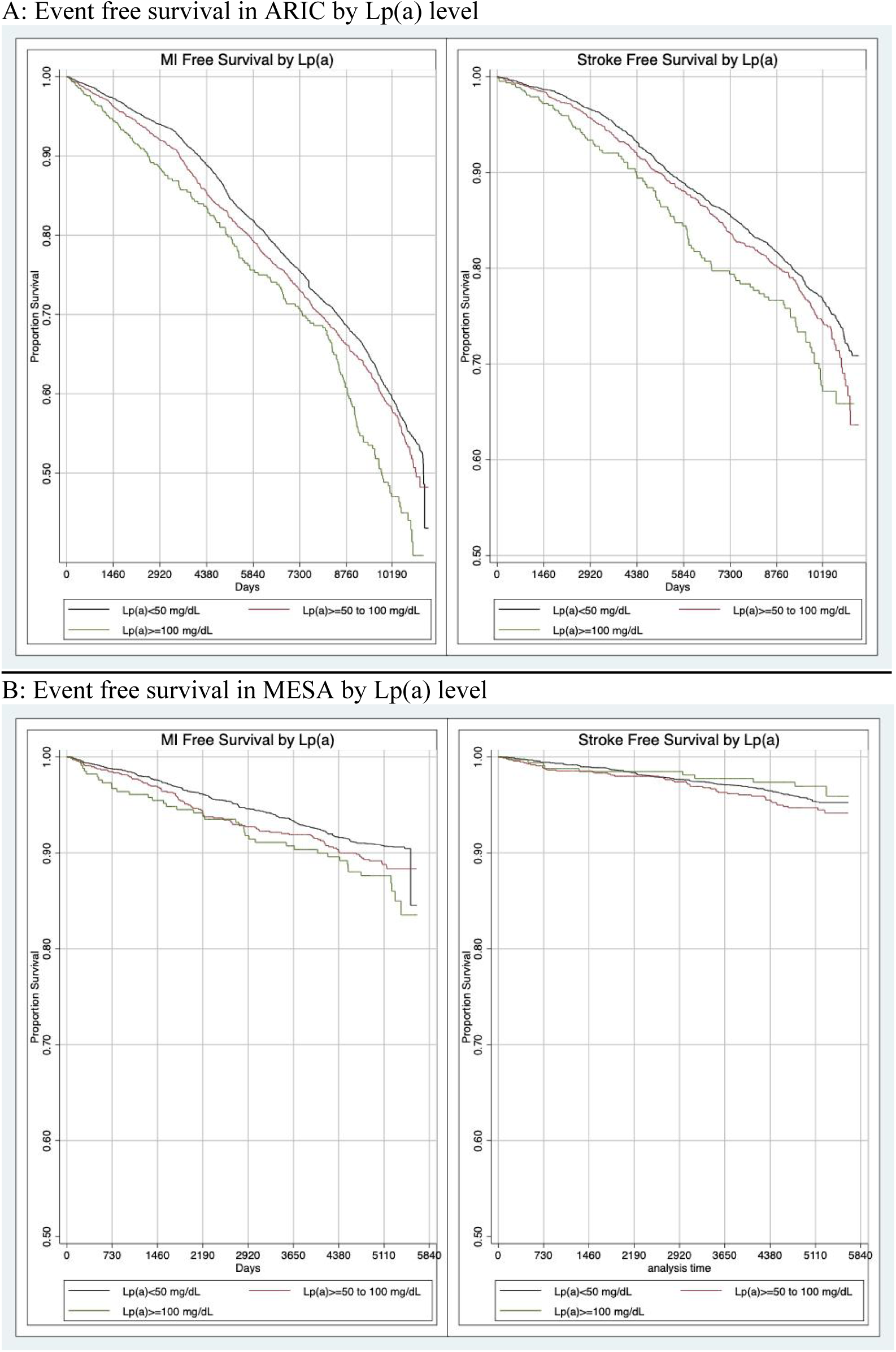
Cox proportional hazards regression results for association of Lp(a) ≥50 mg/dL or <50 mg/dL with myocardial infarction or stroke in ARIC and MESA^a^ ^a^ These proportional hazards regression graphs were generated after exposures (Lp(a) ≥50 mg/dL) were matched using coarsened exact matching to controls (Lp(a) <50 mg/dL). Covariates included age, gender, race and ethnicity, non-HDL-C, SDOH score, and LS7 score.

**Table 5:** Sequential Multivariable Cox Proportional Hazards Regression for Myocardial Infarction in ARIC and MESA After Coarsened Exact Matching.

A: ARIC (n=14,010)
|  | Model 1 |  |  | Model 2 |  |  | Model 3 |  |  |
| --- | --- | --- | --- | --- | --- | --- | --- | --- | --- |
|  | HR | 95% CI | p-value | HR | 95% CI | p-value | HR | 95% CI | p-value |
| Age (per year) | 1.10 | 1.09, 1.10 | <0.001 | 1.09 | 1.09, 1.10 | <0.001 | 1.09 | 1.09, 1.10 | <0.001 |
| Male | 1.54 | 1.44, 1.64 | <0.001 | 1.53 | 1.43, 1.64 | <0.001 | 1.57 | 1.46, 1.68 | <0.001 |
| Race and Ethnicity | - | - | - | - | - | - | - | - | - |
| Non-Hispanic White | Ref | Ref | Ref | Ref | Ref | Ref | Ref | Ref | Ref |
| Non-Hispanic Black | 1.39 | 1.30, 1.49 | <0.001 | 1.17 | 1.09, 1.26 | <0.001 | 0.89 | 0.82, 0.96 | 0.003 |
| Non-HDL-C (per 25 mg/dL increase) | 1.15 | 1.13, 1.17 | <0.001 | 1.14 | 1.12, 1.16 | <0.001 | 1.03 | 1.01, 1.05 | 0.01 |
| Lipoprotein(a) | - | - | - | - | - | - | - | - | - |
| 0 to <50 mg/dL | Ref | Ref | Ref | Ref | Ref | Ref | Ref | Ref | Ref |
| ≥50 to <100 mg/dL | 1.15 | 0.001 | 0.001 | 1.16 | 1.07, 1.27 | 0.001 | 1.13 | 1.04, 1.23 | 0.005 |
| ≥100 mg/dL | 1.53 | <0.001 | <0.001 | 1.53 | 1.33, 1.77 | <0.001 | 1.44 | 1.24, 1.66 | <0.001 |
| SDOH Score (per 1 point increase) | - | - |  | 1.20 | 1.16, 1.24 | <0.001 | 1.12 | 1.08, 1.15 | <0.001 |
| Life's Simple 7 Score (per 1 point increase) | - | - |  | - | - |  | 0.81 | 0.79, 0.82 | <0.001 |

B: MESA (n=5,573)
|  | Model 1 |  |  | Model 2 |  |  | Model 3 |  |  |
| --- | --- | --- | --- | --- | --- | --- | --- | --- | --- |
|  | HR | 95% CI | p-value | HR | 95% CI | p-value | HR | 95% CI | p-value |
| Age (per year) | 1.05 | 1.04, 1.06 | <0.001 | 1.05 | 1.04, 1.06 | <0.001 | 1.04 | 1.03, 1.05 | <0.001 |
| Male | 2.06 | 1.72, 2.46 | <0.001 | 2.24 | 1.86, 2.68 | <0.001 | 2.21 | 1.84, 2.65 | <0.001 |
| Race and Ethnicity | - | - | - | - | - | - | - | - | - |
| Non-Hispanic White | Ref | Ref | Ref | Ref | Ref | Ref | Ref | Ref | Ref |
| Non-Hispanic Black | 1.00 | 0.82, 1.22 | 0.98 | 0.90 | 0.73, 1.11 | 0.32 | 0.77 | 0.62, 0.95 | 0.02 |
| Non-Hispanic Chinese | 0.63 | 0.39, 1.01 | 0.06 | 0.55 | 0.34, 0.90 | 0.02 | 0.63 | 0.38, 1.02 | 0.06 |
| Hispanic | 0.95 | 0.72, 1.25 | 0.70 | 0.72 | 0.53, 0.97 | 0.03 | 0.68 | 0.50, 0.92 | 0.01 |
| Non-HDL-C (per 25 mg/dL increase) | 1.05 | 0.98, 1.12 | 0.18 | 1.05 | 0.98, 1.12 | 0.18 | 0.99 | 0.93, 1.07 | 0.88 |
| Lipoprotein(a) | - | - | - | - | - | - | - | - | - |
| 0 to <50 mg/dL | Ref | Ref | Ref | Ref | Ref | Ref | Ref | Ref | Ref |
| ≥50 to <100 mg/dL | 1.21 | 0.97, 1.52 | 0.09 | 1.23 | 0.98, 1.53 | 0.07 | 1.20 | 0.96, 1.50 | 0.12 |
| ≥100 mg/dL | 1.60 | 1.15, 2.22 | 0.005 | 1.60 | 1.15, 2.23 | 0.005 | 1.57 | 1.13, 2.19 | 0.007 |
| SDOH Score (per 1 point increase) | - | - | - | 1.16 | 1.09, 1.22 | <0.001 | 1.13 | 1.05, 1.17 | <0.001 |
| Life's Simple 7 Score (per 1 point increase) | - | - | - | - | - | - | 0.86 | 0.81, 0.88 | <0.001 |

In fully adjusted models that included SDOH score and LS7 score, Lp(a) >50 mg/dL to <100 mg/dL was associated with stroke in ARIC (1.17, 0.01), but not in MESA (1.18, 0.31) (**Figure 1**). Lp(a) >100 mg/dL was also associated with stroke in ARIC (1.26, 0.02), but not in MESA (0.77, 0.43). Associations between other covariates and MI were similar, including similar shifts in association between race and ethnicity with stroke after adding SDOH and LS7 scores to the model (**Table 6**).

**Table 6:**
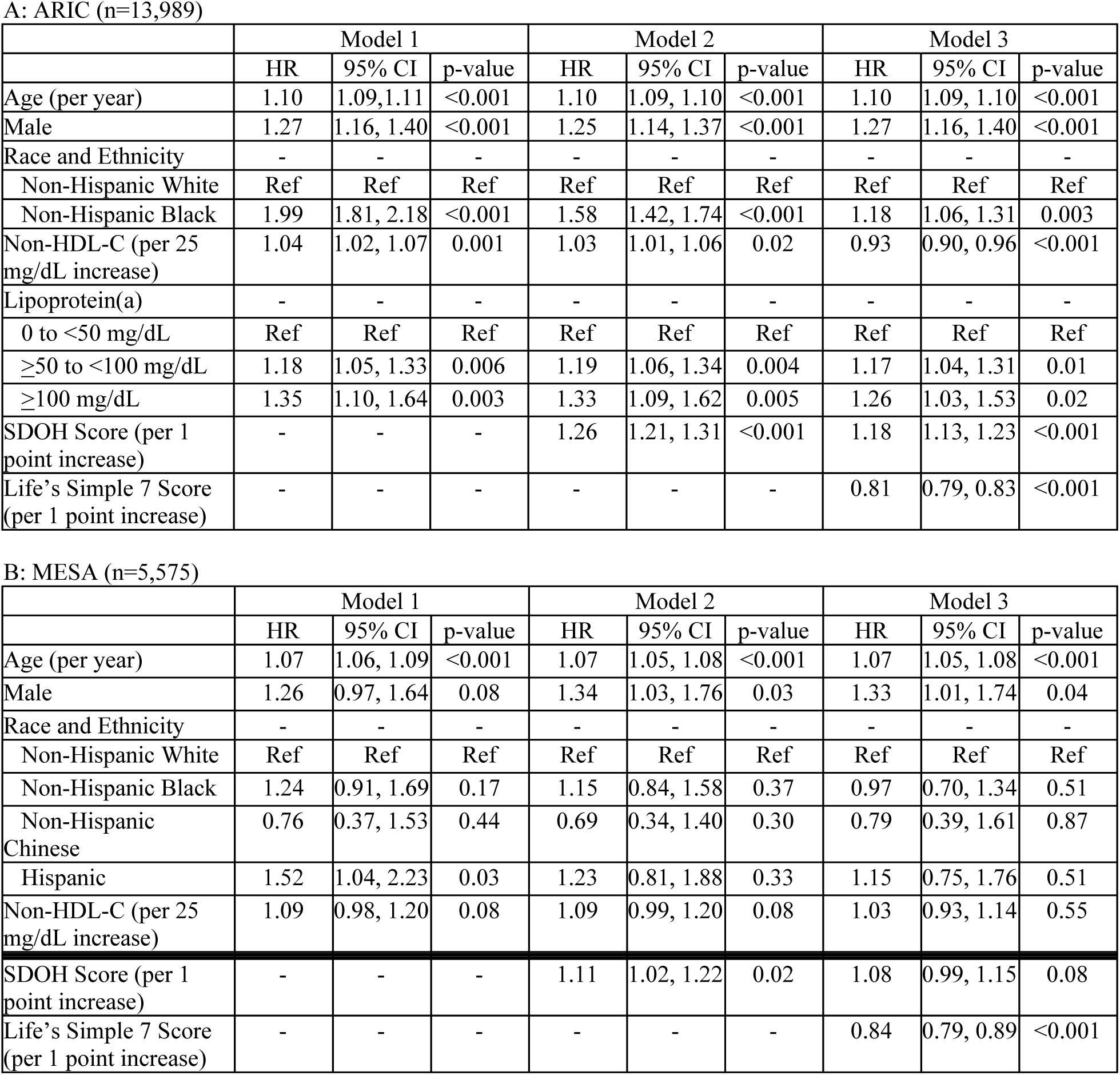
Sequential Multivariable Cox Proportional Hazards Regression for Stroke in ARIC and MESA After Coarsened Exact Matching.

## Discussion

In this observational study of two well-characterized, multi-center US cohorts, Lp(a) was associated with MI in both cohorts and stroke in ARIC but not MESA. Accounting for SDOH and lifestyle factors did not attenuate the association between Lp(a) and MI or stroke. However, lifestyle factors and SDOH were strong predictors of MI or stroke. The large impact of adding SDOH and LS7 scores on the association between race and ethnicity and non-HDL-C with cardiovascular events suggests that SDOH and LS7 may mediate these associations.

Based on the results of two prior studies,^4,5^ we expected that social and lifestyle factors would mediate the risk for ASCVD events related to Lp(a). However, this was not the case since the risk associated with Lp(a) on MI and stroke did not change when accounting for LS7 or SDOH scores. This differed from the ATTICA study since the addition of a Mediterranean diet score led to an abolishment of the risk from Lp(a). This was more similar to the EPIC-Norfolk study wherein despite better lifestyle associated with lower risk but did not abolish the risk among those with elevated Lp(a). Social and lifestyle factors also unlikely moderate the effect of Lp(a) on outcomes since interaction terms were not sufficiently compelling. This was similar to EPIC-Norfolk since interaction tests were also insignificant in this cohort,^4^ but differed from ATTICA wherein an interaction between Mediterranean diet score and Lp(a) was significant (p<0.001).^5^

Lp(a) is an acute phase reactant that may increase in times of inflammation. The chronic stressors from SDOH could have been expected to impact Lp(a) levels.^11^ However, we did not observe that SDOH score correlated with Lp(a) levels. There was however an observed, albeit small impact of LS7 score on Lp(a) levels, which is likely not clinically relevant. This could be because LS7 points are given for lower cholesterol levels, thus there could be collinearity (although post-hoc testing (not shown) did not reveal high collinearity between LS7 or non-HDL-C and Lp(a)). Also, bariatric surgery or lower BMI has correlated in some studies with lower Lp(a) level,^19,20^ although this observation is inconsistent.^21,22^ Lastly, increased healthy lifestyle may could have anti-inflammatory impacts that could impact Lp(a) level.^23^

Even after accounting for many factors, Lp(a) remained associated with MI. This was observed when tested as a linear association and again after matching cases to controls based on Lp(a) as a categorical variable. Our matching analysis achieved excellent balance between cases and controls, with the only unbalanced factors as non-HDL-C, which differed by 3 mg/dL and we do not expect to account for a large difference in associated outcomes. In this matching analysis our results were consistent with prior studies,^1^ Lp(a) ≥50 mg/dL had a moderate effect size on risk for MI, which was higher at levels ≥100 mg/dL. This was similar between cohorts. In a meta-analysis of studies, an Lp(a) of 48 mg/dL had a HR of about 1.1-1.2 and a level of 96 mg/dL or above a HR of about 1.3.^24^ Increased risk at level of ≥50 mg/dL then ≥100 mg/dL were less consistent in the context of stroke wherein Lp(a) is associated with stroke in ARIC but not MESA. Similar to MI in ARIC there was a stepwise increase in effect size at higher Lp(a) levels, which was a lower effect size than that observed in MI. However, the associations in MESA were absent. Prior studies testing the association between Lp(a) and stroke have been variable, with one meta-analysis suggesting that the impact of Lp(a) on increased risk for stroke may be more likely among cohort ≤55 years old.^25^

Our study has important clinical implications in the context that both SDOH and LS7 were consistent predictors of ASCVD. This consistency suggests that SDOH and lifestyle factors should continue to be the focus of clinicians. Health systems should continue to increase screening for SDOH and create pathways for managing SDOH when they are identified.^2,26,27^

There are also implications surrounding the observation that when SDOH and LS7 were added to the model, these mediated the association of other covariates. The associations between non-Hispanic Black or Hispanic participants and CVD events decreased or inverted as SDOH and LS7 were added to the models. This suggests that non-Hispanic Black or Hispanic participants’ SDOH and LS7 scores were confounding associations as mediators between race and ethnicity with MI or stroke. Addressing SDOH and lifestyle factors can therefore be seen as a chance for healthy equity in managing risk for ASCVD.^2,26,28^ Future studies looking at associations between race and ethnicity and events should recognize our observations and that differences across race and ethnicity are driven by non-biologic factors (e.g., structural inequities).^8,29,30^

Lastly, when SDOH and lifestyle factors were included in the models, the effect size for the association between non-HDL-C and outcomes was reduced or eliminated. The largest shift occurred when the LS7 score was added to the model. This suggests that some of the association between atherogenic lipids and risk for ASCVD is driven by lifestyle factors. The LS7 score includes a lipid component, thus there could be collinearity of variables in the model. Lastly, the effect size of non-HDL-C on outcomes could have been reduced in the context that we were unable to track therapies introduced over time. If those with higher non-HDL-C were more likely to be treated with lipid lowering therapies then this could confound this association.

### Strengths

A strength of this study is that we used two large cohorts with long follow up periods and many of the observations were similar between cohorts. Additionally we used multiple methods and yielded similar results, including matching cases of Lp(a) ≥50 mg/dL to controls that were similar on many factors.

### Limitations

Our study has limitations. There may be additional SDOH that were not measured in either cohort. Several of the items are self-reported, which could be subject to social desirability bias and incomplete data collection. Lastly, these data are observational and further work needs to be done to understand the mechanisms by which SDOH and lifestyle alter the biochemistry of Lp(a).

### Conclusions

Our findings support that Lp(a) is an independent risk factor for cardiovascular events and that this associated risk is not greatly impacted by SDOH or lifestyle factors. In addition, SDOH and lifestyle factors are strong predictors of ASCVD risk that may explain some of the association between race and ethnicity or non-HDL-C with MI or stroke. This impact of SDOH and LS7 should be considered when designing and interpreting observational and clinical studies.

## Data Availability

Data are available for access by request to NIH BioLINCC

https://biolincc.nhlbi.nih.gov/studies/mesa/

https://biolincc.nhlbi.nih.gov/studies/aric/

## Acknowledgements

None

## References

1. Tsimikas S. A Test in Context: Lipoprotein(a): Diagnosis, Prognosis, Controversies, and Emerging Therapies. J Am Coll Cardiol [Internet]. 2017;69:692–711. Available from: 10.1016/j.jacc.2016.11.042

2. Brandt EJ, Tobb K, Cambron JC, Ferdinand K, Douglass P, Nguyen PK, Vijayaraghavan K, Islam S, Thamman R, Rahman S, Pendyal A, Sareen N, Yong C, Palaniappan L, Ibebuogu U, Tran A, Bacong AM, Lundberg G, Watson K. Assessing and Addressing Social Determinants of Cardiovascular Health: JACC State-of-the-Art Review. J Am Coll Cardiol [Internet]. 2023;81:1368– 1385. Available from: 10.1016/j.jacc.2023.01.042

3. Lloyd-Jones DM, Hong Y, Labarthe D, Mozaffarian D, Appel LJ, Van Horn L, Greenlund K, Daniels S, Nichol G, Tomaselli GF, Arnett DK, Fonarow GC, Ho PM, Lauer MS, Masoudi FA, Robertson RM, Roger V, Schwamm LH, Sorlie P, Yancy CW, Rosamond WD, American Heart Association Strategic Planning Task Force and Statistics Committee. Defining and setting national goals for cardiovascular health promotion and disease reduction: the American Heart Association’s strategic Impact Goal through 2020 and beyond. *Circulation* [Internet]. 2010;121:586–613. Available from: 10.1161/CIRCULATIONAHA.109.192703

4. Perrot N, Verbeek R, Sandhu M, Boekholdt SM, Hovingh GK, Wareham NJ, Khaw KT, Arsenault BJ. Ideal cardiovascular health influences cardiovascular disease risk associated with high lipoprotein(a) levels and genotype: The EPIC-Norfolk prospective population study. Atherosclerosis [Internet]. 2017;256:47–52. Available from: 10.1016/j.atherosclerosis.2016.11.010

5. Foscolou A, Georgousopoulou E, Magriplis E, Naumovski N, Rallidis L, Matalas AL, Chrysohoou C, Tousoulis D, Pitsavos C, Panagiotakos D. The mediating role of Mediterranean diet on the association between Lp(a) levels and cardiovascular disease risk: A 10-year follow-up of the ATTICA study. Clin Biochem [Internet]. 2018;60:33–37. Available from: 10.1016/j.clinbiochem.2018.07.011

6. Satcher D. Include a social determinants of health approach to reduce health inequities. Public Health Rep [Internet]. 2010;125 Suppl 4:6–7. Available from: 10.1177/00333549101250s402

7. Palacio A, Mansi R, Seo D, Suarez M, Garay S, Medina H, Tang F, Tamariz L. Social determinants of health score: does it help identify those at higher cardiovascular risk? Am J Manag Care [Internet]. 2020;26:e312–e318. Available from: 10.37765/ajmc.2020.88504

8. Bundy JD, Mills KT, He H, LaVeist TA, Ferdinand KC, Chen J, He J. Social determinants of health and premature death among adults in the USA from 1999 to 2018: a national cohort study. Lancet Public Health [Internet]. 2023;8:e422–e431. Available from: 10.1016/S2468-2667(23)00081-6

9. Teshale AB, Htun HL, Owen A, Gasevic D, Phyo AZZ, Fancourt D, Ryan J, Steptoe A, Freak-Poli R. The Role of Social Determinants of Health in Cardiovascular Diseases: An Umbrella Review. J Am Heart Assoc [Internet]. 2023;12:e029765. Available from: 10.1161/JAHA.123.029765

10. Havranek EP, Mujahid MS, Barr DA, Blair IV, Cohen MS, Cruz-Flores S, Davey-Smith G, Dennison-Himmelfarb CR, Lauer MS, Lockwood DW, Rosal M, Yancy CW, American Heart Association Council on Quality of Care and Outcomes Research, Council on Epidemiology and Prevention, Council on Cardiovascular and Stroke Nursing, Council on Lifestyle and Cardiometabolic Health, and Stroke Council. Social Determinants of Risk and Outcomes for Cardiovascular Disease: A Scientific Statement From the American Heart Association. Circulation [Internet]. 2015;132:873–898. Available from: 10.1161/CIR.0000000000000228

11. Feingold KR, Grunfeld C. The Effect of Inflammation and Infection on Lipids and Lipoproteins [Internet]. MDText.com, Inc.; 2022 [cited 2023 Jul 11]. Available from: https://www.ncbi.nlm.nih.gov/books/NBK326741/

12. Gaubatz JW, Heideman C, Gotto AM Jr, Morrisett JD, Dahlen GH. Human plasma lipoprotein [a]. Structural properties. J Biol Chem [Internet]. 1983;258:4582–4589. Available from: https://www.ncbi.nlm.nih.gov/pubmed/6220008

13. Gaubatz JW, Cushing GL, Morrisett JD. Quantitation, isolation, and characterization of human lipoprotein (a). Methods Enzymol. 1986;129:167–186.

14. Virani SS, Brautbar A, Davis BC, Nambi V, Hoogeveen RC, Sharrett AR, Coresh J, Mosley TH, Morrisett JD, Catellier DJ, Folsom AR, Boerwinkle E, Ballantyne CM. Associations between lipoprotein(a) levels and cardiovascular outcomes in black and white subjects: the Atherosclerosis Risk in Communities (ARIC) Study. Circulation [Internet]. 2012;125:241–249. Available from: 10.1161/circulationaha.111.045120

15. Agarwala A, Pokharel Y, Saeed A, Sun W, Virani SS, Nambi V, Ndumele C, Shahar E, Heiss G, Boerwinkle E, Konety S, Hoogeveen RC, Ballantyne CM. The association of lipoprotein(a) with incident heart failure hospitalization: Atherosclerosis Risk in Communities study. Atherosclerosis [Internet]. 2017;262:131–137. Available from: 10.1016/j.atherosclerosis.2017.05.014

16. Guan W, Cao J, Steffen BT, Post WS, Stein JH, Tattersall MC, Kaufman JD, McConnell JP, Hoefner DM, Warnick R, Tsai MY. Race is a key variable in assigning lipoprotein(a) cutoff values for coronary heart disease risk assessment: the Multi-Ethnic Study of Atherosclerosis. Arterioscler Thromb Vasc Biol [Internet]. 2015;35:996–1001. Available from: 10.1161/atvbaha.114.304785

17. Rikhi R, Hammoud A, Ashburn N, Snavely AC, Michos ED, Chevli P, Tsai MY, Herrington D, Shapiro MD. Relationship of low-density lipoprotein-cholesterol and lipoprotein(a) to cardiovascular risk: The Multi-Ethnic Study of Atherosclerosis (MESA). Atherosclerosis [Internet]. 2022;363:102–108. Available from: 10.1016/j.atherosclerosis.2022.10.004

18. Tsimikas S, Karwatowska-Prokopczuk E, Gouni-Berthold I, Tardif J-C, Baum SJ, Steinhagen-Thiessen E, Shapiro MD, Stroes ES, Moriarty PM, Nordestgaard BG, Xia S, Guerriero J, Viney NJ, O’Dea L, Witztum JL. Lipoprotein(a) Reduction in Persons with Cardiovascular Disease. N Engl J Med [Internet]. 2020;382:244–255. Available from: https://www.nejm.org/doi/full/10.1056/NEJMoa1905239

19. Jamialahmadi T, Reiner Ž, Alidadi M, Kroh M, Almahmeed W, Ruscica M, Sirtori C, Rizzo M, Santos RD, Sahebkar A. The Effect of Bariatric Surgery on Circulating Levels of Lipoprotein (a): A Meta-analysis. Biomed Res Int [Internet]. 2022;2022:8435133. Available from: 10.1155/2022/8435133

20. Hoursalas A, Tsarouhas K, Tsitsimpikou C, Kolovou G, Vardavas A, Hoursalas I, Spandidos DA, Milionis H, Elisaf M, Tsiara S. Moderately elevated lipoprotein (a) levels are associated with an earlier need for percutaneous coronary intervention in recurrent cardiovascular disease. Exp Ther Med [Internet]. 2022;24:444. Available from: 10.3892/etm.2022.11371

21. Corsetti JP, Sterry JA, Sparks JD, Sparks CE, Weintraub M. Effect of weight loss on serum lipoprotein(a) concentrations in an obese population. Clin Chem [Internet]. 1991;37:1191–1195. Available from: https://www.ncbi.nlm.nih.gov/pubmed/1830250

22. Berk KA, Yahya R, Verhoeven AJM, Touw J, Leijten FP, van Rossum EF, Wester VL, Lips MA, Pijl H, Timman R, Erhart G, Kronenberg F, Roeters van Lennep JE, Sijbrands EJG, Mulder MT. Effect of diet-induced weight loss on lipoprotein(a) levels in obese individuals with and without type 2 diabetes. Diabetologia [Internet]. 2017;60:989–997. Available from: 10.1007/s00125-017-4246-y

23. Kim S, Chang Y, Cho J, Hong YS, Zhao D, Kang J, Jung H-S, Yun KE, Guallar E, Ryu S, Shin H. Life’s Simple 7 Cardiovascular Health Metrics and Progression of Coronary Artery Calcium in a Low-Risk Population. Arterioscler Thromb Vasc Biol [Internet]. 2019;39:826–833. Available from: 10.1161/ATVBAHA.118.311821

24. Erqou S, Kaptoge S, Perry PL, Di Angelantonio E, Thompson A, White IR, Marcovina SM, Collins R, Thompson SG, Danesh J. Lipoprotein(a) concentration and the risk of coronary heart disease, stroke, and nonvascular mortality. JAMA [Internet]. 2009;302:412–423. Available from: 10.1001/jama.2009.1063

25. Nave AH, Lange KS, Leonards CO, Siegerink B, Doehner W, Landmesser U, Steinhagen-Thiessen E, Endres M, Ebinger M. Lipoprotein (a) as a risk factor for ischemic stroke: a meta-analysis. Atherosclerosis [Internet]. 2015;242:496–503. Available from: 10.1016/j.atherosclerosis.2015.08.021

26. Velarde G, Bravo-Jaimes K, Brandt EJ, Wang D, Douglass P, Castellanos LR, Rodriguez F, Palaniappan L, Ibebuogu U, Bond R, Ferdinand K, Lundberg G, Thaman R, Vijayaraghavan K, Watson K. Locking the Revolving Door: Racial Disparities in Cardiovascular Disease. J Am Heart Assoc [Internet]. 2023;e025271. Available from: 10.1161/JAHA.122.025271

27. Vanjani R, Reddy N, Giron N, Bai E, Martino S, Smith M, Harrington-Steppen S, Trimbur MC. The Social Determinants of Health — Moving Beyond Screen-and-Refer to Intervention. N Engl J Med [Internet]. 2023;389:569–573. Available from: 10.1056/NEJMms2211450

28. Arnett DK, Blumenthal RS, Albert MA, Buroker AB, Goldberger ZD, Hahn EJ, Himmelfarb CD, Khera A, Lloyd-Jones D, McEvoy JW, Michos ED, Miedema MD, Munoz D, Smith SC Jr, Virani SS, Williams KA Sr, Yeboah J, Ziaeian B. 2019 ACC/AHA Guideline on the Primary Prevention of Cardiovascular Disease: Executive Summary: A Report of the American College of Cardiology/American Heart Association Task Force on Clinical Practice Guidelines. J Am Coll Cardiol [Internet]. 2019;140:e563–e595. Available from: 10.1016/j.jacc.2019.03.009

29. Braveman P, Parker Dominguez T. Abandon “Race.” Focus on Racism. Front Public Health [Internet]. 2021;9:689462. Available from: 10.3389/fpubh.2021.689462

30. Javed Z, Haisum Maqsood M, Yahya T, Amin Z, Acquah I, Valero-Elizondo J, Andrieni J, Dubey P, Jackson RK, Daffin MA, Cainzos-Achirica M, Hyder AA, Nasir K. Race, Racism, and Cardiovascular Health: Applying a Social Determinants of Health Framework to Racial/Ethnic Disparities in Cardiovascular Disease. Circ Cardiovasc Qual Outcomes [Internet]. 2022;15:e007917. Available from: 10.1161/CIRCOUTCOMES.121.007917

